# Psychometric development and validation of the Hospital Resilience Index

**DOI:** 10.1101/2023.03.16.23287371

**Authors:** Philipp Lirk, Haroon Janjua, Michael Rogers, Stephen Flynn, Paul Kuo, Ricardo Pietrobon

**Author notes:** **Address for correspondence** Dr. Philipp Lirk, Department of Anesthesiology, Perioperative and Pain Medicine, Brigham and Women’s Hospital, Harvard Medical School, 75 Francis Street, Boston MA 02115, USA.

## Abstract

**Importance:** Throughout the past decades, the United States healthcare system has seen increased efforts to promote resilience to external or internal stressors. However, measuring outcomes remains a challenge, mainly because most indices rely on markers that need to be gathered from different data sources. An alternative would be to use readily available large-scale data. The American Hospital Association (AHA) annual survey gathers over 1,000 data points from more than 6,000 hospitals. The RAND corporation provides information on the financial health and viability of hospitals and health care systems.

**Objective:** Our study aimed to establish and psychometrically validate a new Hospital Resilience Index.

**Design:** We took the two databases as primary data sources, and defined hospital closure as the main negative outcome. We performed descriptive statistics, and regression analysis of the databases.

**Main outcomes and Measures:** Likelihood of hospital closure.

**Results:** Our findings show that a combination of eleven variables is strongly associated with the likelihood of hospital closure. These factors mirror smaller hospital size, lack of ancillary functions, staffing structure of the hospital, size of facilities, number of surgeries performed, Medicare discharges, operating expenses, and medical school affiliation as a teaching hospital. We further classified hospitals with a low HRI (<25) or high HRI (>25). In this setup, we found that both hospitals with a low HRI and hospitals that would subsequently close were characterized by smaller patient census, smaller numbers of surgeries, fewer beds, a smaller staff, and a lower operating margin.

**Conclusions and Relevance:** Together, these factors would point to a higher resilience to external stressors in larger, more expanded hospitals and healthcare systems, offering a broad range of services, and having a higher operating margin. The higher tiers of the healthcare system therefore seem to have more resilience, but the recent Covid-19 pandemic exhibited how much all tiers in the system are needed to respond to extraordinary crises such as pandemics or large natural disasters. Future research should seek to determine whether tracking the HRI over time may be a tool to identify hospitals at risk of closure.

**Key points:** *Question:* Can existing databases (e.g., American Hospital Association) be used to quantify resilience of a hospital, and predict closure?

*Findings:* In this study, we combined data from the American Hospital Association Database, and the RAND corporation database to create a Hospital Resiliency Index, with the hard outcome of hospital closure. A combination of eleven variables characterized hospitals which would subsequently close, mostly pointing towards smaller, rural hospitals.

*Meaning:* Our study is the basis for potential monitoring of hospital resilience using data readily available for every U.S. hospital on an annual basis, to allow for identification of at-risk hospitals over time.

## Introduction

Over the past 20 years, the United States healthcare system has put increasing focus on Resilience, which encompasses a process of anticipating and planning for potential disasters, the readiness to respond and recover as efficiently as possible, and the inherent ability of the system to learn from the event to inform future standards of operation.^1,2^

Several metrics have been proposed to objectively assess aspects of resilience and preparedness in the Health Care sector. On one end of the spectrum, there are general frameworks such as the eleven essential resilience elements proposed by Khan.^3^ On the other end of the spectrum are detailed and comprehensive assessments such as the resilience indices proposed by Dobalian,^4^ Kruk,^5^ Meyer,^6^ and McCarthy.^7^ The downside of these metrics, however, is that the data needed for assessments are not readily available, rather they need to be aggregated individually from different sources in each site investigated. Automated data retrieval based on existing databases would seem the more prudent strategy, e.g., the Hospital Medical Surge Preparedness Index (HMSPI).^8, 9^

We propose here to expand the concept of the HMSPI to generate a Hospital Resilience Index (HRI), linking AHA survey and RAND corporation datasets. Our objective is to develop and psychometrically assess the Hospital Resilience Index for hospitals and healthcare facilities across the United States in relation to their capacity to function efficiently during stressful events such as mass casualty events, natural disasters or pandemics.

## Methods

### Study design

HRI was created based on our preliminary work related to hospital preparedness.^8, 9^ Our study used data from 2018 to 2020 from the American Hospital Association (AHA) annual survey ^10^ and RAND (research and development) Hospital Data.^11^

### Ethics

The University of South Florida Institutional Review Board evaluated this study and considered it exempt.

### Setting and hospitals

We based HRI on two primary data sources, accessed through Wharton Research Data Services (WRDS), wrds.wharton.upenn.edu, accession date 2022-04-21:

#### 1. American Hospital Association (AHA)

The AHA annual survey is a comprehensive dataset for analyzing and comparing healthcare industry trends among community hospitals in the United States. The database enables analysis of trends in utilization, personnel, revenues, and expenses across local, regional, and national markets,^10^ inlcusing over 1,000 data fields for over 6,300 hospitals, with a response rate of more than 75% from all hospitals in the country. A detailed report of hospital closures in 2019, the most recent year available, was separately purchased from AHA.

#### 2. RAND Hospital Data

The RAND Hospital Data is a tool compiled by the RAND Corporation, a research organization committed to developing solutions to public policy concerns to improve community safety, security, health, and prosperity worldwide.^11^ The RAND Hospital Data tool was designed to widen the user base and simplify analytical tasks for persons dealing with data daily, e.g., through the Healthcare Cost Report Information System (HCRIS). We linked both datasets through Medicare hospital identifiers for the years 2018-2020, 2020 being the latest publicly available year at this point.

To evaluate the association between HRI and price compliance, we used a list that indicates 326 hospitals with price compliance in 47 US states.

### Hospital Resilience Index (HRI)

We used the AHA survey from 2018 to 2020 ^10^ linked to RAND Hospital Data.^11^ Item selection was driven by a conceptual framework for understanding hospital resilience.^12^ This framework combines different stages associated with resilience (preparedness, response, and recovery) and hospital characteristics (staff, infrastructure, management, and logistics) that influence hospital resilience.^12^ We convened a group of psychometric, clinical, and resilience experts to select the most likely items to be a part of the final index. We then submitted these items to the psychometric analyses reported below, including item correlation analysis with graphical heatmaps, scree plots, and exploratory and confirmatory factor analyses.

### Items

We initially evaluated variables from the AHA survey in the development of the HRI characterizing the hospital (size, revenue, worker base), and its place in the community (outreach, ambulatory surgery centers, ambulance services, cooperation with nursing homes), summarized in eTable 1. Regarding RAND Hospital Data, we focused on hospital size, revenue, operating expenses, administrative costs, and margins.

**Table 1.** Summary of factor loadings for the selected variables.

| **Variable** | **Factor 1** | **h<sup>2</sup>** |
| --- | --- | --- |
| Full-time registered nurses (categorized) | 0.975 | 0.951 |
| Adjusted patient days (categorized) | 0.882 | 0.777 |
| Number of Operating Rooms (categorized) | 0.943 | 0.89 |
| Total facility expenses (categorized) | 0.985 | 0.97 |
| Total facility employee benefits (categorized) | 0.959 | 0.92 |
| Inpatient surgical operations (categorized) | 0.959 | 0.919 |
| Total facility Medicaid discharges (categorized) | 0.943 | 0.89 |
| Total facility admissions (categorized) | 0.985 | 0.971 |
| Operating expenses (categorized) | 0.979 | 0.959 |
| Total Full-time hospital unit personnel (categorized) | 0.976 | 0.953 |
| Total facility Medicare discharges (categorized) | 0.967 | 0.934 |

The detailed statistical analysis is described in the eSupplement.

## Results

### Exploratory analysis

In our demographic analysis of the 5,924 hospitals in the survey (eTable 1), we found that 60% of the hospitals were located in the South region, followed by Midwest (19%), West (11.1%), and Northeast (9.52%). A total of 6.35% were owned wholly or partially by physicians or a group of physicians, and 11.1% were part of a Medical school affiliation reported to the AMA. Out of 5,924 hospitals, 63 closed in 2019, and 5,861 continued operations. The first level of analysis was to identify factors that showed a significant difference between operating and closed hospitals. In total, thirty-eight key metrics were observed. In general, smaller hospitals that offered less services made up the bulk of closures. For example, 64% of the closed hospitals provided <=15 medical and surgical adult beds, compared to 26.9% of the hospitals that did not close in 2019. Hospitals containing an emergency department represented 19% of closed hospitals, contrasting to 60.2% of hospitals that continued active. Certified trauma centers were present in only 1.59% of the closed hospitals group, whereas this variable represented 28.8% of the hospitals continuing operations. Closed hospitals also had very few admissions, 50% of closed hospitals had less than 1,828 admissions per year (averaging 5 or less per day), as compared to approximately 25% in operating hospitals. The same applied for hospital administration and costs, closed hospieals were, on average, much smaller than “surviving” ones. A significant percentage of hospitals that did not close in 2019 presented community health education initiatives (57.9%), in contrast to only 17.5% in the case of those that closed in the same year. We observed the same for hospitals engaged in health screenings, which presented a great difference in percentages (55.6% vs. 14.3%, respectively).

### Exploratory factor analysis (EFA)

We next performed a series of exploratory factor analyses including the 38 items from the survey. First, we evaluated the correlation between studied variables through a correlation plot (eFigure 1). Highly positively correlated variables included the number of admissions, total facility inpatient days, surgical operations, Medicare and Medicaid discharges, facility expenses, employee benefits, full-time or part-time registered nurses/equivalent, and full-time unit personnel, general medical and adult beds, the total number of beds, number of operating rooms, total net expenses and operating expenses. The operating margin was demonstrated to be highly correlated to the total margin, and the same was verified for Medicare inpatient and outpatient margins in relation to Medicare inpatient margin. The part-time number of registered nurses was positively correlated with the total part-time unit personnel. Full-time and part-time physicians and dentists were also highly correlated to full-time equivalent physicians and dentists.

**Figure 1.**
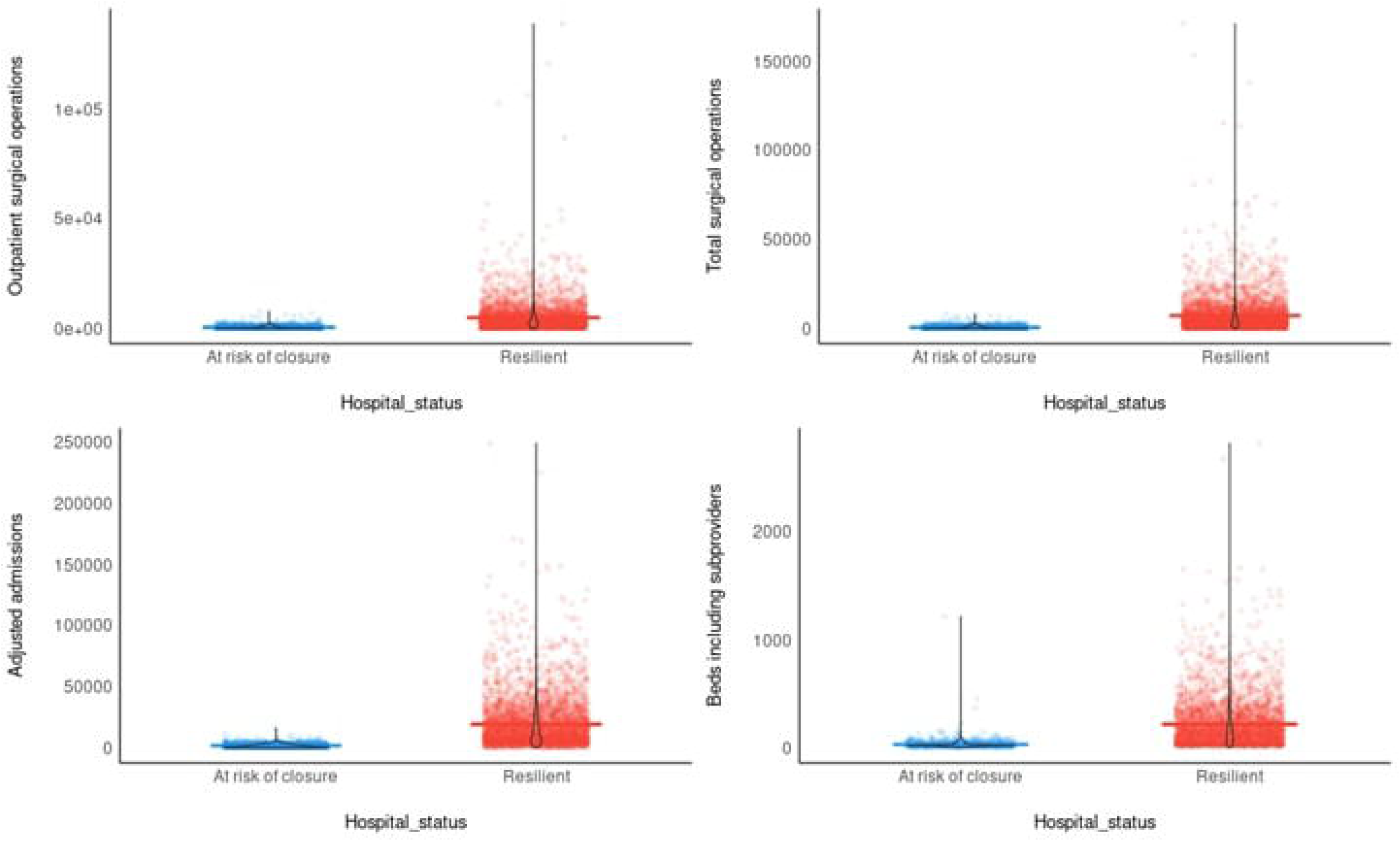
Comparison between hospitals with a Hospital Resilience Index (HRI) above vs. below the overall mean HRI (25). We present hospitals with an HRI above 25 as ‘resilient’ and those below that level as a ‘at risk of closure.’

Rural health clinics were negatively correlated or lacked correlation to almost all other variables, as seen for the FIPS State Code and Medicare inpatient/outpatient margin. Health screenings, community health education, and the presence of certified trauma centers were weakly correlated to operating and total margins.

Our factor loading analysis identified that the best fit occurred using a one-factor structure solution, in which there is no particular item concentration but a high structural fit of the grouped items, as seen in the MAP analysis. We report only the items with a factor loading above 0.3 (eTable 2). Our results show that these items are well-fitted to the structure, confirmed by Factor 1 scores all above 0.8. In the same way, the EFA diagram shows Factor 1 dominating the subjected items, confirmed by the arrows pointing in the same direction (eFigure 2).

**Table 2.** Internal reliability

| Cronbach's alpha | Raykov's<br>omega | Bentler's<br>omega | McDonald's<br>omega | Average<br>variance (AVE) |
| --- | --- | --- | --- | --- |
| 0.979 | 0.979 | 0.979 | 0.98 | 0.81 |

**Figure 2.**
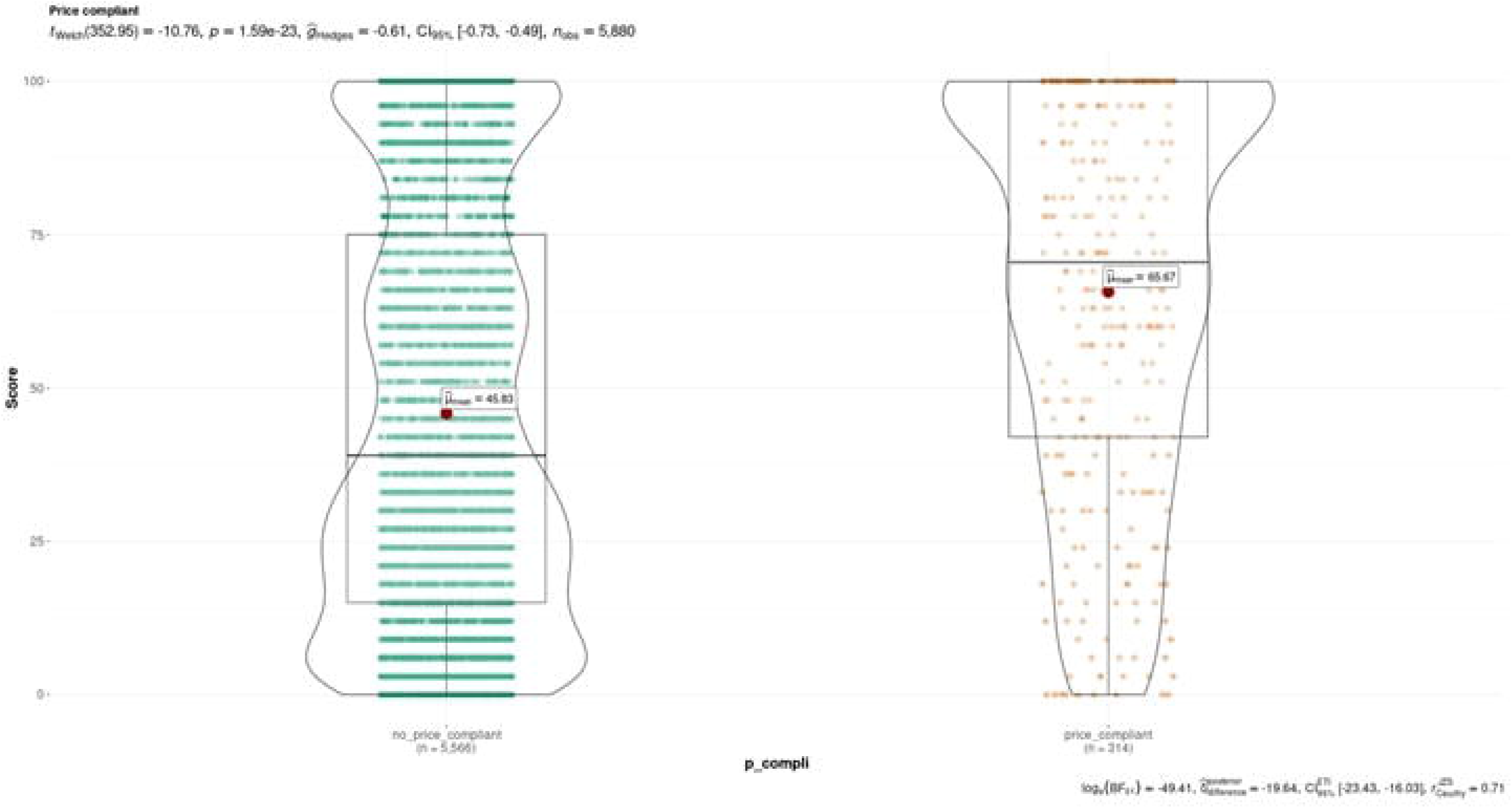
Comparison between Hospital Resilience Index for hospitals with vs. without price compliance in 2019.

We analyzed a variety of fit statistics to determine the model fitness for the confirmatory factor analysis (Table 1). From the CFI, NFI, and TLI metrics, we confirmed the model fitness is acceptable (values > 0.9). The RMSEA and SRMR metrics also corroborate with the previous validation, in which lower scores are expected for best model fitness. The internal validity analysis also demonstrated high reliability in our index, represented by scores higher than 0.9 in the metrics analyzed (Table 2).

We next classified hospitals with an HRI above 25 as ‘resilient’ and those below that level as ‘at risk of closure’ in relation to the variables outpatient surgical operation, total surgical operations, adjusted admissions, and beds, including sub providers. This result indicates that hospitals with an HRI > 25 for these variables present a lower risk of closure (Figure 1).

When comparing HRI for hospitals with or without price compliance in 2019 (Figure 2), we identified a significant statistical difference between hospitals that provide these data and a higher HRI (P < 0.001) and those that do not provide price compliance and a lower HRI (p < 0.001).

We attempted to identify which variables were contributing to a higher or lower extent to HRI and the probability of closure in 2019. In eFigures 3 and 4, we discriminate the contribution of each item from HRI to the predicted probability of closure for exemplary hospitals that did close in 2019.

Subsequently, we tested our resilience index by evaluating the HRI for Venice Regional Bayfront Health over three consecutive years in order to obtain an indication of its possible closure, which occurred later in 2022.{Ellison, 2022 #4780} Overall, the HRI indicates that the hospital had an ongoing decline in the index from 2018 to 2020 (eFigure 5).

We next evaluated if there was a difference in HRI when considering the years before closing in 2020 (eFigure 6). When analyzed by group, hospitals that would close in that year presented a median HRI in 2018 and 2019 of 24, whereas hospitals that would not close exhibited a median HRI of 42 (p<0.001). The HRI of institutions that kept active was substantially higher than that of closed hospitals in 2018 and 2019. In 2020, the index maintained a similar pattern for active hospitals.

## Discussion

The current study sought to establish the Hospital Resilience Index (HRI) as a measure of hospital resilience under duress, with hospital closures chosen as the outcome to psychometrically evaluate our proposed metric. Our main findings are that a combination of 11 variables is strongly associated with the likelihood of hospital closure, which we take to be the ultimate manifestation of failure to exhibit resilience.

Prediction of hospital closure has previously been suggested, based on financial metrics, ^17, 18^ for example derived from the Centers for Medicaid and Medicare Services (CMS) Healthcare Cost Report Information System (HCRIS).^18^ However, the financial aspect alone may not fully capture the overall resilience of the system. What would therefore be needed is a synthesis of financial, administrative and medical data,^19^ with data readily available for extraction, such that analyses can be adapted quickly, and trends over time assembled. The latter aspect is especially important since hospital closures are often foreshadowed ahead of time.^20^ We chose to combine data from the American Hospital Association (AHA) annual survey with RAND corporation data to paint a comprehensive picture of the resilience of American hospitals, and to investigate whether factors amenable to data extraction from these databases would predict hospital closure.

In our analysis, we first looked at the basic statistics of the included hospitals, and found that 11% of hospitals in the AHA survey reported an affiliation with a medical school. One factor which quickly became apparent was that hospital scope and size were important factors influencing potential closure. For example, hospitals that would go on to close had a very low likelihood of maintaining or being associated with a nursing home type long-term care facility (1.6% versus 64%), had a significantly higher likelihood of employing less than 40 full-time nursing staff, performed smaller numbers of surgeries, and were twice as likely to have less than 40 patients on average in house on a daily basis. Another highly interesting factor was that community outreach, in the form of health education initiatives, was performed by almost 60% of open hospitals, but only by 17.5% of hospitals that went on to close. This could be interpreted as a potential harbinger of impending financial difficulty, which often leads to cuts in services to restore financial stability.^21^ A negative operating margin was observed in more than 70% of closed hospitals, but only 24% of open hospitals, with our analysis predating the significant strain on hospitals during and after the first two and a half years of the pandemic.^21^ Moreover, the trends observed by us are reflected one level lower, on the service level, where closure of emergency departments show similar trends, with closed EDs showing smaller patient census, and low profit margins.^22^ The items identified here are also reflective of other measures of financial distress, for example those determining the modified Altman Z-score.^17^

As a second step, we performed a series of exploratory analyses including the statistics listed above, in total 38 variables. We could quantify the correlation between the 38 items and hospital closure (eFigure 1) and distill the list of factors down to 11, characterized by a factor loading above 0.3 (Table 2). These variables encompass both the staffing structure of the hospital, size of facilities, number of surgeries performed, Medicare discharges, and operating expenses.

The resulting risk score, the HRI presented here, uses a heuristic approach to classify hospitals according to their resilience. One more complex and, perhaps, accurate approach would be to employ artificial intelligence (AI) algorithms to predict closures, as they have been employed to predict hospital treatment effects ^23^ and improve operations.^24^ Given that true machine learning algorithms are not in use currently, we aimed to implement a more pragmatic, albeit highly structured, development methodology. In doing so, we need to realize that a risk score is not deterministic, but clearly signal an increase in the likelihood of an adverse event, such as closure. In our study, the HRI was highly significantly associated with the risk of closure, but it was not a guarantee of that outcome. Rather than being used in an isolated manner, we propose to further develop this index to be included in comprehensive assessments of hospital operations, as part of a composite metric. Another potential use for the HRI is the possibility to track hospitals over time, as decreasing financial freedom and service interruptions may be early harbingers of impending collapse.^18^

Even though the risk score was constructed using regression analysis, the variables found do reflect important policy discussions. First, smaller hospitals are more likely to close than large ones. Hospital closures have accelerated throughout the last decade,^19^ with rural hospitals accounting for almost 80% of closures.^25^ Closing hospitals has two sides, one is that redirecting patients to other facilities may improve efficiency and financial outcomes,^26^ but for patients and communities, hospital closures are primarily associated with decreased access to care, and again the main effect is felt most in rural populations,^27^ especially the elderly and poor.^28, 29^ Conversely, healthcare facilities have substantial positive economic benefits, both in terms of revenue flowing into a community, and by promoting sustainable, well-paying jobs, contributing to a stable labor market.^30^ The effects of hospital closure are partly attenuated when the former hospital is converted to another type of facility, such as an ambulatory center.^31^ It follows that prediction of impending hospital failure would be an important asset.^18^

Second is the question whether hospital closure or failure to comply with regulations would necessarily indicate poor management. As an example from this study, when correlating the HRI with price compliance, the obligation of hospitals to transparently share the cost of their treatments ahead of time, we found that price compliant hospitals featured a much higher median HRI (65) than price incompliant ones (42, P < 0.001). This could be interpreted in two ways. On one hand, general management quality, including compliance, could be interpreted as suboptimal in closing hospitals, but on the other hand, it has been suggested that efforts that go beyond pure patient care may be difficult to achieve for smaller hospitals that are already struggling.^32^ Difficulty in maintenance of surgical volume may be related to workforce shortages.^33^

Third, it is important to consider the implications of our study in terms of the distribution of resilience across the system. Both in general resilience to stressors, and in detailed preparedness efforts to be able to deal with mass casualty events, the emphasis has been on the preservation of these function in large medical centers. However, both the experience from Superstorm Sandy (Flynn, 2015) and the Covid-19 pandemic showed that when large-scale disaster strikes, all tiers of the healthcare system are needed for the overall system to cope.^34, 35^ The majority of Americans receive care in community healthcare centers, especially minority and impoverished communities,^35^ but resilience and preparedness efforts are confined often to larger Institutions.^34^ When major disasters hit, however, influx from non-resilient, smaller, units may overwhelm even the capacity of major hospitals considered the backbone of modern healthcare provision.^34^

Finally, limitations of the present study should be addressed. First, our data analysis only covered a three-year period, and we believe that surveillance over longer periods of time would allow for a more detailed interpretation of the performance, and second, our HRI does not indicate causality, but the same can be said about any modeling approach. Nevertheless, we believe that the current model is a solid basis on which to pursue further research, focused on the interaction between financial and medical indicators and quality of care, as well as focusing on social circumstances of hospital closures.

In conclusion, we describe here the HRI as a composite marker of hospital performance, and evaluate its usefulness in the detection of impending hospital closure. We describe 11 markers, strongly associated with hospital closure. Future research should focus on the integration of the HRI into a composite hospital evaluation, and its association with quality of care and social determinants of health.

## Supporting information

Supplemental Methodology

eTables

eFigures legends

eFigure 1

eFigure 2

eFigure 3a

eFigure 3b

eFigure 4a

eFigure 4b

eFigure 5

eFigure 6

## Data Availability

Data analysis plans are available, data itself is from a database (AHA/RAND) that requires purchase of the data, so individual data will not be available.

