## Supplemental Methodology for "Psychometric development and validation of the Hospital Resilience Index"

**Statistical analysis**

**Exploratory analysis.** We began our exploratory analysis by visually evaluating all categorical variables (e.g., information on whether the hospital restricts admissions primarily to children, whether the hospital is owned in whole or in part by physicians or a physician group, and type of hospital) to determine their frequency, percentage, near-zero variation, missing value patterns, as well as distribution for numeric variables (e.g., number of beds, number of full-time physicians and dentists, and number of surgical operations).^13^ When a categorical variable has a low frequency of distinct values over the sample size, it is said to have near-zero variance, implying that the variable is almost constant. We addressed near-zero variance by merging variable categorizations. We also applied variable transformations, like logarithmic and Box-Cox, when numeric variables did not present a normal distribution. To address missing variables, we used imputation algorithms, followed by sensitivity analyses, to ensure that our results were stable with or without imputation.^14^ We utilized standardized mean difference (SMD) for the exploratory analysis, consisting of the proportions or mean difference divided by the pooled standard deviation. We applied the following guidelines when interpreting SMD magnitude: (1) A minor effect as SMD = 0.2; (2) a medium effect as SMD = 0.5; and (3) a large effect as SMD = 0.8.^15^ We also included p values for t-tests (for numeric variables) and Chi-square tests (for categorical variables). We determined the statistical significance of p-values as those below 0.05.

**Assessment of hospital closures.** We conducted the initial analysis using data from 2018, whereas hospital closure information was obtained from 2019 using the detailed report on hospital closures supplemental to the AHA database.

**Exploratory factor analysis.** First, we used correlation matrices and plots as exploratory analysis tools to better understand the association across all items.^15^ We also conducted a series of exploratory factor analyses using oblique and orthogonal rotations to explore different factorial solutions underlying the data, using maximum likelihood as the extraction method. Our heuristic for selecting factor solutions included: (a) scree plots, (b) solutions that were theoretically justifiable, and (c) solutions where items loaded with values above 0.30 on a single factor while all other loadings were below that threshold.

**Confirmatory factor analysis.** Next, we conducted a confirmatory factor analysis. Our model was theoretically justified fitting statistics for confirmatory factor analyses, which included the following goodness-of-fit measures: fit function value, chi-squared value based on the fit function, p-value for chi-squared value and degrees of freedom, chi-squared value for baseline model, degrees of freedom for baseline model, p-value for the baseline model, comparative fit index (CFI), Tucker-Lewis index (TLI), non-normed fit index (NNFI), relative fit index (RFI), normed fit index (NFI), parsimony normed fit index (PNFI), incremental fit index (IFI), relative noncentrality index (RNI), logarithm of the likelihood statistic, logarithm of the likelihood statistic for baseline model, akaike information criterion (AIC), bayesian information criterion (BIC), bayesian information criterion adjusted for sample size (BICc), root mean square error of approximation (RMSEA), p-value associated with hypothesis of RMSEA < 0.05, root mean square residual (includes means) (RMR), root mean square residual (no means), standardized root mean square residual (includes means) (SRMR), Bentler standardized root mean square residual (includes means), Bentler standardized root mean square residual (no means), Bollen standardized root mean square residual (includes means), Bollen standardized root mean square residual (no means), MPlus standardized root mean square residual (includes means), MPlus Standardized Root Mean Square Residual (no means), critical n for alpha = 0.05, critical n for alpha = 0.01, goodness-of-fit index (GFI), adjusted goodness-of-fit index, parsimony goodness-of-fit index (PGFI), mcdonald fit index (MFI), and expected cross-validation index.

**Internal reliability.** Normalized scores went from 0 to 100 and were then used for reliability assessment through Cronbach’s alpha and Omega ^16^ within each factor.^15^ Score normalization was conducted for each item, then each subdomain, and then the overall HRI score, using a standard formula

xnormalized=x−xmin/xmax−mmin

Since there are no maximum upper values for numeric variables, we normalized the score for the maximum value of each variable for the year 2020.

**Validity.** We assessed validity by comparing HRI scores in 2019 against a hospital closure the following year (2020). We also evaluated the association between HRI and price compliant-hospitals. Comparisons were established using logistic regression models calculated with 95% confidence intervals.
