## Supplementary material for "Psychometric development and validation of the Hospital Resilience Index": eTables

### Supplemental Materials

### eTable 1

Variables description and comparison between hospitals undergoing or not undergoing a closure in 2019.

| **Variable [Missing]** | **Total (5,924)** | **Closed in 2019 (63)** | **Not closed in 2019 (5,861)** | **p** |
| --- | --- | --- | --- | --- |
| Does the hospital restrict admissions primarily to children? [1,634] | 89 (1.5%) | 0 (0%) | 89 (1.52%) | p = 0.979 (Chi-square test) |
| Bed size code [0] |  |  |  | p = 0.053 (Chi-square test) |
| -6-24 beds | 795 (13.4%) | 15 (23.8%) | 780 (13.3%) |  |
| -100-199 beds | 1,175 (19.8%) | 10 (15.9%) | 1,165 (19.9%) |  |
| -400-499 beds | 183 (3.09%) | 1 (1.59%) | 182 (3.11%) |  |
| -200-299 beds | 605 (10.2%) | 7 (11.1%) | 598 (10.2%) |  |
| -50-99 beds | 1,127 (19%) | 15 (23.8%) | 1,112 (19%) |  |
| -25-49 beds | 1,390 (23.5%) | 15 (23.8%) | 1,375 (23.5%) |  |
| -300-399 beds | 340 (5.74%) | 0 (0%) | 340 (5.8%) |  |
| -500 or more beds | 309 (5.22%) | 0 (0%) | 309 (5.27%) |  |
| Is hospital owned in whole or in part by physicians or a physician group? [1,634] | 223 (3.76%) | 4 (6.35%) | 219 (3.74%) | p = 0.047 (Chi-square test) |
| FIPS State code [0] |  |  |  | p = 0.038 (Chi-square test) |
| -Others | 77 (1.3%) | 0 (0%) | 77 (1.31%) |  |
| -Northeast | 751 (12.7%) | 6 (9.52%) | 745 (12.7%) |  |
| -South | 2,434 (41.1%) | 38 (60.3%) | 2,396 (40.9%) |  |
| -Midwest | 1,632 (27.5%) | 12 (19%) | 1,620 (27.6%) |  |
| -West | 1,030 (17.4%) | 7 (11.1%) | 1,023 (17.5%) |  |
| Medical school affiliation reported to AMA [0] | 1,648 (27.8%) | 7 (11.1%) | 1,641 (28%) | p = 0.005 (Chi-square test) |
| Member of Council of Teaching Hospital of the Association of American Medical Colleges (COTH) [0] | 263 (4.44%) | 1 (1.59%) | 262 (4.47%) | p = 0.425 (Chi-square test) |
| Medicare certification by the U.S. Department of Health and Human Services [4] | 5,785 (97.7%) | 60 (95.2%) | 5,725 (97.7%) | p = 0.367 (Chi-square test) |
| Rural Referral Center [0] | 319 (5.38%) | 0 (0%) | 319 (5.44%) | p = 0.105 (Chi-square test) |
| Does the hospital maintain a separate nursing home type of long-term care unit? [4] | 499 (8.42%) | 1 (1.59%) | 498 (8.5%) | p = 0.082 (Chi-square test) |
| General medical and surgical (adult) beds (categorized) [1,634] |  |  |  | p < 0.001 (Chi-square test) |
| -<= 15 | 1,165 (19.7%) | 16 (64%) | 1,149 (26.9%) |  |
| -<= 31 | 982 (16.6%) | 2 (8%) | 980 (23%) |  |
| -<= 108 | 1,075 (18.1%) | 5 (20%) | 1,070 (25.1%) |  |
| -> 108 | 1,068 (18%) | 2 (8%) | 1,066 (25%) |  |
| Total hospital beds (calculated) (categorized) [4] |  |  |  | p = 0.027 (Chi-square test) |
| -<= 30 | 1,527 (25.8%) | 24 (38.1%) | 1,503 (25.7%) |  |
| -<= 80 | 1,456 (24.6%) | 19 (30.2%) | 1,437 (24.5%) |  |
| -<= 194 | 1,458 (24.6%) | 12 (19%) | 1,446 (24.7%) |  |
| -> 194 | 1,479 (25%) | 8 (12.7%) | 1,471 (25.1%) |  |
| Ambulatory surgery center - hospital [1,634] | 1,069 (18%) | 3 (4.76%) | 1,066 (18.2%) | p = 0.206 (Chi-square test) |
| Ambulatory surgery center - health system [1,634] | 784 (13.2%) | 0 (0%) | 784 (13.4%) | p = 0.035 (Chi-square test) |
| Ambulatory surgery center - joint venture [1,634] | 272 (4.59%) | 1 (1.59%) | 271 (4.62%) | p = 0.944 (Chi-square test) |
| Community outreach - hospital [1,634] | 3,051 (51.5%) | 10 (15.9%) | 3,041 (51.9%) | p = 0.001 (Chi-square test) |
| Community outreach - health system [1,634] | 737 (12.4%) | 0 (0%) | 737 (12.6%) | p = 0.044 (Chi-square test) |
| Community outreach - joint venture [1,634] | 54 (0.912%) | 1 (1.59%) | 53 (0.904%) | p = 0.739 (Chi-square test) |
| Emergency Department - hospital [1,634] | 3,543 (59.8%) | 12 (19%) | 3,531 (60.2%) | p < 0.001 (Chi-square test) |
| Emergency Department - health system [1,634] | 571 (9.64%) | 0 (0%) | 571 (9.74%) | p = 0.095 (Chi-square test) |
| Emergency Department - joint venture [1,634] | 77 (1.3%) | 1 (1.59%) | 76 (1.3%) | p = 0.938 (Chi-square test) |
| Certified trauma center - hospital [1,634] | 1,690 (28.5%) | 1 (1.59%) | 1,689 (28.8%) | p < 0.001 (Chi-square test) |
| Certified trauma center - health system [1,634] | 662 (11.2%) | 0 (0%) | 662 (11.3%) | p = 0.062 (Chi-square test) |
| Certified trauma center - joint venture [1,634] | 62 (1.05%) | 1 (1.59%) | 61 (1.04%) | p = 0.816 (Chi-square test) |
| Community Health Education - hospital [1,634] | 3,407 (57.5%) | 11 (17.5%) | 3,396 (57.9%) | p < 0.001 (Chi-square test) |
| Community Health Education - health system [1,634] | 742 (12.5%) | 1 (1.59%) | 741 (12.6%) | p = 0.134 (Chi-square test) |
| Community Health Education - joint venture [1,634] | 55 (0.928%) | 1 (1.59%) | 54 (0.921%) | p = 0.749 (Chi-square test) |
| Health screenings - hospital [1,634] | 3,269 (55.2%) | 9 (14.3%) | 3,260 (55.6%) | p < 0.001 (Chi-square test) |
| Health screenings - health system [1,634] | 737 (12.4%) | 1 (1.59%) | 736 (12.6%) | p = 0.137 (Chi-square test) |
| Health screenings - joint venture [1,634] | 49 (0.827%) | 2 (3.17%) | 47 (0.802%) | p = 0.022 (Chi-square test) |
| Rural health clinic - hospital [1,634] | 1,016 (17.2%) | 0 (0%) | 1,016 (17.3%) | p = 0.011 (Chi-square test) |
| Rural health clinic - health system [1,634] | 441 (7.44%) | 2 (3.17%) | 439 (7.49%) | p = 0.963 (Chi-square test) |
| Rural health clinic - joint venture [1,634] | 64 (1.08%) | 1 (1.59%) | 63 (1.07%) | p = 0.834 (Chi-square test) |
| Full-time physicians and dentists (categorized) [4] |  |  |  | p = 0.01 (Chi-square test) |
| -<= 0 | 1,946 (32.8%) | 27 (42.9%) | 1,919 (32.8%) |  |
| -<= 3 | 1,249 (21.1%) | 16 (25.4%) | 1,233 (21.1%) |  |
| -<= 11 | 1,281 (21.6%) | 16 (25.4%) | 1,265 (21.6%) |  |
| -> 11 | 1,444 (24.4%) | 4 (6.35%) | 1,440 (24.6%) |  |
| Full-time registered nurses (categorized) [0] |  |  |  | p < 0.001 (Chi-square test) |
| -<= 30 | 1,521 (25.7%) | 33 (52.4%) | 1,488 (25.4%) |  |
| -<= 69 | 1,450 (24.5%) | 12 (19%) | 1,438 (24.5%) |  |
| -<= 243 | 1,472 (24.8%) | 13 (20.6%) | 1,459 (24.9%) |  |
| -> 243 | 1,481 (25%) | 5 (7.94%) | 1,476 (25.2%) |  |
| Part-time physicians and dentists (categorized) [0] |  |  |  | p = 0.01 (Chi-square test) |
| -<= 0 | 2,624 (44.3%) | 31 (49.2%) | 2,593 (44.2%) |  |
| -<= 1 | 1,023 (17.3%) | 18 (28.6%) | 1,005 (17.1%) |  |
| -<= 3 | 917 (15.5%) | 9 (14.3%) | 908 (15.5%) |  |
| -> 3 | 1,360 (23%) | 5 (7.94%) | 1,355 (23.1%) |  |
| Part-time registered nurses (categorized) [0] |  |  |  | p = 0.002 (Chi-square test) |
| -<= 14 | 1,562 (26.4%) | 25 (39.7%) | 1,537 (26.2%) |  |
| -<= 34 | 1,444 (24.4%) | 18 (28.6%) | 1,426 (24.3%) |  |
| -<= 95 | 1,438 (24.3%) | 17 (27%) | 1,421 (24.2%) |  |
| -> 95 | 1,480 (25%) | 3 (4.76%) | 1,477 (25.2%) |  |
| Full time equivalent physicians and dentists (categorized) [0] |  |  |  | p = 0.017 (Chi-square test) |
| -<= 0 | 1,828 (30.9%) | 26 (41.3%) | 1,802 (30.7%) |  |
| -<= 4 | 1,403 (23.7%) | 18 (28.6%) | 1,385 (23.6%) |  |
| -<= 13 | 1,227 (20.7%) | 14 (22.2%) | 1,213 (20.7%) |  |
| -> 13 | 1,466 (24.7%) | 5 (7.94%) | 1,461 (24.9%) |  |
| Full time equivalent registered nurses (categorized) [0] |  |  |  | p < 0.001 (Chi-square test) |
| -<= 40 | 1,532 (25.9%) | 31 (49.2%) | 1,501 (25.6%) |  |
| -<= 88 | 1,432 (24.2%) | 14 (22.2%) | 1,418 (24.2%) |  |
| -<= 306 | 1,482 (25%) | 13 (20.6%) | 1,469 (25.1%) |  |
| -> 306 | 1,478 (24.9%) | 5 (7.94%) | 1,473 (25.1%) |  |
| (Estimation Flag) Surgical Operations Inpatient [0] |  |  |  | p < 0.001 (Chi-square test) |
| -Estimated | 2,006 (33.9%) | 42 (66.7%) | 1,964 (33.5%) |  |
| -Reported | 3,894 (65.7%) | 20 (31.7%) | 3,874 (66.1%) |  |
| -Expanded | 24 (0.405%) | 1 (1.59%) | 23 (0.392%) |  |
| (Estimation Flag) Surgical Operations Outpatient [0] |  |  |  | p < 0.001 (Chi-square test) |
| -Estimated | 2,006 (33.9%) | 42 (66.7%) | 1,964 (33.5%) |  |
| -Reported | 3,894 (65.7%) | 20 (31.7%) | 3,874 (66.1%) |  |
| -Expanded | 24 (0.405%) | 1 (1.59%) | 23 (0.392%) |  |
| (Estimation Flag) Surgical Operations Total [0] |  |  |  | p < 0.001 (Chi-square test) |
| -Estimated | 2,010 (33.9%) | 42 (66.7%) | 1,968 (33.6%) |  |
| -Reported | 3,890 (65.7%) | 20 (31.7%) | 3,870 (66%) |  |
| -Expanded | 24 (0.405%) | 1 (1.59%) | 23 (0.392%) |  |
| Inpatient surgical operations (categorized) [0] |  |  |  | p < 0.001 (Chi-square test) |
| -<= 5 | 1,487 (25.1%) | 29 (46%) | 1,458 (24.9%) |  |
| -<= 356 | 1,475 (24.9%) | 16 (25.4%) | 1,459 (24.9%) |  |
| -<= 1,789 | 1,482 (25%) | 13 (20.6%) | 1,469 (25.1%) |  |
| -> 1,789 | 1,480 (25%) | 5 (7.94%) | 1,475 (25.2%) |  |
| Outpatient surgical operations (categorized) [0] |  |  |  | p < 0.001 (Chi-square test) |
| -<= 185 | 1,482 (25%) | 30 (47.6%) | 1,452 (24.8%) |  |
| -<= 1,622 | 1,480 (25%) | 14 (22.2%) | 1,466 (25%) |  |
| -<= 4,158 | 1,481 (25%) | 15 (23.8%) | 1,466 (25%) |  |
| -> 4,158 | 1,481 (25%) | 4 (6.35%) | 1,477 (25.2%) |  |
| Total surgical operations (categorized) [0] |  |  |  | p < 0.001 (Chi-square test) |
| -<= 218 | 1,481 (25%) | 30 (47.6%) | 1,451 (24.8%) |  |
| -<= 2,064 | 1,481 (25%) | 15 (23.8%) | 1,466 (25%) |  |
| -<= 6,138 | 1,482 (25%) | 12 (19%) | 1,470 (25.1%) |  |
| -> 6,138 | 1,480 (25%) | 6 (9.52%) | 1,474 (25.1%) |  |
| Adjusted patient days (categorized) [0] |  |  |  | p < 0.001 (Chi-square test) |
| -<= 14,478 | 1,481 (25%) | 31 (49.2%) | 1,450 (24.7%) |  |
| -<= 37,524 | 1,481 (25%) | 16 (25.4%) | 1,465 (25%) |  |
| -<= 94,460 | 1,481 (25%) | 14 (22.2%) | 1,467 (25%) |  |
| -> 94,460 | 1,481 (25%) | 2 (3.17%) | 1,479 (25.2%) |  |
| Number of Operating Rooms (categorized) [1,824] |  |  |  | p = 0.026 (Chi-square test) |
| -<= 2 | 1,458 (24.6%) | 14 (66.7%) | 1,444 (35.4%) |  |
| -<= 5 | 741 (12.5%) | 1 (4.76%) | 740 (18.1%) |  |
| -<= 11 | 879 (14.8%) | 3 (14.3%) | 876 (21.5%) |  |
| -> 11 | 1,022 (17.3%) | 3 (14.3%) | 1,019 (25%) |  |
| Adjusted admissions (categorized) [0] |  |  |  | p < 0.001 (Chi-square test) |
| -<= 1,828 | 1,481 (25%) | 29 (46%) | 1,452 (24.8%) |  |
| -<= 5,758 | 1,481 (25%) | 20 (31.7%) | 1,461 (24.9%) |  |
| -<= 17,580 | 1,481 (25%) | 12 (19%) | 1,469 (25.1%) |  |
| -> 17,580 | 1,481 (25%) | 2 (3.17%) | 1,479 (25.2%) |  |
| Total facility admissions (categorized) [0] |  |  |  | p < 0.001 (Chi-square test) |
| -<= 579 | 1,482 (25%) | 30 (47.6%) | 1,452 (24.8%) |  |
| -<= 1,954 | 1,480 (25%) | 16 (25.4%) | 1,464 (25%) |  |
| -<= 7,846 | 1,481 (25%) | 12 (19%) | 1,469 (25.1%) |  |
| -> 7,846 | 1,481 (25%) | 5 (7.94%) | 1,476 (25.2%) |  |
| Total facility Medicare discharges (categorized) [4] |  |  |  | p < 0.001 (Chi-square test) |
| -<= 283 | 1,481 (25%) | 24 (38.1%) | 1,457 (24.9%) |  |
| -<= 841 | 1,481 (25%) | 24 (38.1%) | 1,457 (24.9%) |  |
| -<= 3,708 | 1,478 (24.9%) | 10 (15.9%) | 1,468 (25.1%) |  |
| -> 3,708 | 1,480 (25%) | 5 (7.94%) | 1,475 (25.2%) |  |
| Total facility Medicare days (categorized) [0] |  |  |  | p = 0.091 (Chi-square test) |
| -<= 1,808 | 1,481 (25%) | 16 (25.4%) | 1,465 (25%) |  |
| -<= 6,338 | 1,481 (25%) | 22 (34.9%) | 1,459 (24.9%) |  |
| -<= 19,489 | 1,481 (25%) | 17 (27%) | 1,464 (25%) |  |
| -> 19,489 | 1,481 (25%) | 8 (12.7%) | 1,473 (25.1%) |  |
| Total facility Medicaid discharges (categorized) [0] |  |  |  | p = 0.012 (Chi-square test) |
| -<= 42 | 1,490 (25.2%) | 26 (41.3%) | 1,464 (25%) |  |
| -<= 338 | 1,472 (24.8%) | 16 (25.4%) | 1,456 (24.8%) |  |
| -<= 1,493 | 1,481 (25%) | 13 (20.6%) | 1,468 (25%) |  |
| -> 1,493 | 1,481 (25%) | 8 (12.7%) | 1,473 (25.1%) |  |
| Total facility Medicaid days (categorized) [0] |  |  |  | p = 0.023 (Chi-square test) |
| -<= 271 | 1,483 (25%) | 25 (39.7%) | 1,458 (24.9%) |  |
| -<= 2,359 | 1,479 (25%) | 14 (22.2%) | 1,465 (25%) |  |
| -<= 9,672 | 1,481 (25%) | 16 (25.4%) | 1,465 (25%) |  |
| -> 9,672 | 1,481 (25%) | 8 (12.7%) | 1,473 (25.1%) |  |
| Total Part-time hospital unit personnel (categorized) [0] |  |  |  | p = 0.002 (Chi-square test) |
| -<= 50 | 1,503 (25.4%) | 24 (38.1%) | 1,479 (25.2%) |  |
| -<= 113 | 1,470 (24.8%) | 18 (28.6%) | 1,452 (24.8%) |  |
| -<= 296 | 1,470 (24.8%) | 18 (28.6%) | 1,452 (24.8%) |  |
| -> 296 | 1,481 (25%) | 3 (4.76%) | 1,478 (25.2%) |  |
| Total Full-time hospital unit personnel (categorized) [0] |  |  |  | p < 0.001 (Chi-square test) |
| -<= 130 | 1,486 (25.1%) | 36 (57.1%) | 1,450 (24.7%) |  |
| -<= 292 | 1,481 (25%) | 8 (12.7%) | 1,473 (25.1%) |  |
| -<= 840 | 1,477 (24.9%) | 17 (27%) | 1,460 (24.9%) |  |
| -> 840 | 1,480 (25%) | 2 (3.17%) | 1,478 (25.2%) |  |
| Total facility expenses (categorized) [0] |  |  |  | p < 0.001 (Chi-square test) |
| -<= 21,965,622 | 1,481 (25%) | 30 (47.6%) | 1,451 (24.8%) |  |
| -<= 55,993,888 | 1,481 (25%) | 16 (25.4%) | 1,465 (25%) |  |
| -<= 183,426,252 | 1,481 (25%) | 14 (22.2%) | 1,467 (25%) |  |
| -> 183,426,252 | 1,481 (25%) | 3 (4.76%) | 1,478 (25.2%) |  |
| Total facility inpatient days (categorized) [0] |  |  |  | p = 0.007 (Chi-square test) |
| -<= 4,678 | 1,481 (25%) | 20 (31.7%) | 1,461 (24.9%) |  |
| -<= 16,476 | 1,487 (25.1%) | 24 (38.1%) | 1,463 (25%) |  |
| -<= 43,969 | 1,475 (24.9%) | 13 (20.6%) | 1,462 (24.9%) |  |
| -> 43,969 | 1,481 (25%) | 6 (9.52%) | 1,475 (25.2%) |  |
| Total facility employee benefits (categorized) [0] |  |  |  | p < 0.001 (Chi-square test) |
| -<= 2,103,603 | 1,481 (25%) | 35 (55.6%) | 1,446 (24.7%) |  |
| -<= 5,607,305 | 1,481 (25%) | 13 (20.6%) | 1,468 (25%) |  |
| -<= 16,967,004 | 1,481 (25%) | 13 (20.6%) | 1,468 (25%) |  |
| -> 16,967,004 | 1,481 (25%) | 2 (3.17%) | 1,479 (25.2%) |  |
| Beds (categorized) [1,341] |  |  |  | p = 0.12 (Chi-square test) |
| -<= 25 | 1,525 (25.7%) | 7 (25%) | 1,518 (33.3%) |  |
| -<= 74 | 778 (13.1%) | 9 (32.1%) | 769 (16.9%) |  |
| -<= 199 | 1,137 (19.2%) | 8 (28.6%) | 1,129 (24.8%) |  |
| -> 199 | 1,143 (19.3%) | 4 (14.3%) | 1,139 (25%) |  |
| Beds including subproviders (categorized) [1,338] |  |  |  | p = 0.362 (Chi-square test) |
| -<= 25 | 1,156 (19.5%) | 6 (21.4%) | 1,150 (25.2%) |  |
| -<= 97 | 1,143 (19.3%) | 11 (39.3%) | 1,132 (24.8%) |  |
| -<= 228 | 1,142 (19.3%) | 6 (21.4%) | 1,136 (24.9%) |  |
| -> 228 | 1,145 (19.3%) | 5 (17.9%) | 1,140 (25%) |  |
| Inpatient length of stay (categorized) [1,345] |  |  |  | p = 0.632 (Chi-square test) |
| -<= 3.68 | 1,145 (19.3%) | 10 (35.7%) | 1,135 (24.9%) |  |
| -<= 4.36 | 1,145 (19.3%) | 6 (21.4%) | 1,139 (25%) |  |
| -<= 5.35 | 1,144 (19.3%) | 6 (21.4%) | 1,138 (25%) |  |
| -> 5.35 | 1,145 (19.3%) | 6 (21.4%) | 1,139 (25%) |  |
| Major teaching hospital interns and residents to bed [1,330] | 370 (6.25%) | 1 (1.59%) | 369 (6.3%) | p = 0.599 (Chi-square test) |
| Minor teaching hospital interns and residents to bed [1,330] | 861 (14.5%) | 5 (7.94%) | 856 (14.6%) | p = 1 (Chi-square test) |
| Type of hospital [1,330] |  |  |  |  |
| -General short term | 4,504 (76%) | 28 (100%) | 4,476 (98%) |  |
| -General long term | 2 (0.034%) | 0 (0%) | 2 (0.044%) |  |
| -Cancer | 11 (0.186%) | 0 (0%) | 11 (0.241%) |  |
| -Psych | 0 (0%) | 0 (0%) | 0 (0%) |  |
| -Rehabilitation | 1 (0.017%) | 0 (0%) | 1 (0.022%) |  |
| -Religious Non Medical | 0 (0%) | 0 (0%) | 0 (0%) |  |
| -Childrens | 0 (0%) | 0 (0%) | 0 (0%) |  |
| -Alcohol and drugs | 0 (0%) | 0 (0%) | 0 (0%) |  |
| -Other | 76 (1.28%) | 0 (0%) | 76 (1.66%) |  |
| Operating margin (categorized) [1,441] |  |  |  | p < 0.001 (Chi-square test) |
| -<= -0.042 | 1,121 (18.9%) | 20 (71.4%) | 1,101 (24.7%) |  |
| -<= 0.021 | 1,121 (18.9%) | 4 (14.3%) | 1,117 (25.1%) |  |
| -<= 0.084 | 1,120 (18.9%) | 3 (10.7%) | 1,117 (25.1%) |  |
| -> 0.084 | 1,121 (18.9%) | 1 (3.57%) | 1,120 (25.1%) |  |
| Total unreimbursed and Uncompensated care Medicaid and SCHIP cost (categorized) [1,404] |  |  |  | p = 0.097 (Chi-square test) |
| -<= 1,776,506 | 1,130 (19.1%) | 12 (42.9%) | 1,118 (24.9%) |  |
| -<= 5,506,551 | 1,130 (19.1%) | 8 (28.6%) | 1,122 (25%) |  |
| -<= 14,813,120 | 1,130 (19.1%) | 4 (14.3%) | 1,126 (25.1%) |  |
| -> 14,813,120 | 1,130 (19.1%) | 4 (14.3%) | 1,126 (25.1%) |  |
| Administrative costs (categorized) [1,337] |  |  |  | p = 0.291 (Chi-square test) |
| -<= 0.169 | 1,147 (19.4%) | 5 (17.9%) | 1,142 (25%) |  |
| -<= 0.201 | 1,147 (19.4%) | 4 (14.3%) | 1,143 (25.1%) |  |
| -<= 0.238 | 1,146 (19.3%) | 9 (32.1%) | 1,137 (24.9%) |  |
| -> 0.238 | 1,147 (19.4%) | 10 (35.7%) | 1,137 (24.9%) |  |
| Administrative costs including share of mixed categories (categorized) [1,338] |  |  |  | p = 0.097 (Chi-square test) |
| -<= 0.223 | 1,147 (19.4%) | 4 (14.3%) | 1,143 (25.1%) |  |
| -<= 0.26 | 1,146 (19.3%) | 4 (14.3%) | 1,142 (25.1%) |  |
| -<= 0.305 | 1,146 (19.3%) | 8 (28.6%) | 1,138 (25%) |  |
| -> 0.305 | 1,147 (19.4%) | 12 (42.9%) | 1,135 (24.9%) |  |
| Medicare inpatient day share (categorized) [1,348] |  |  |  | p = 0.291 (Chi-square test) |
| -<= 0.296 | 1,144 (19.3%) | 10 (35.7%) | 1,134 (24.9%) |  |
| -<= 0.404 | 1,144 (19.3%) | 4 (14.3%) | 1,140 (25.1%) |  |
| -<= 0.539 | 1,144 (19.3%) | 9 (32.1%) | 1,135 (25%) |  |
| -> 0.539 | 1,144 (19.3%) | 5 (17.9%) | 1,139 (25%) |  |
| Medicaid inpatient day share (categorized) [1,670] |  |  |  | p = 0.927 (Chi-square test) |
| -<= 0.024 | 1,064 (18%) | 6 (23.1%) | 1,058 (25%) |  |
| -<= 0.056 | 1,063 (17.9%) | 6 (23.1%) | 1,057 (25%) |  |
| -<= 0.117 | 1,063 (17.9%) | 6 (23.1%) | 1,057 (25%) |  |
| -> 0.117 | 1,064 (18%) | 8 (30.8%) | 1,056 (25%) |  |
| Non Medicare non Medicaid inpatient day share (categorized) [1,672] |  |  |  | p = 0.57 (Chi-square test) |
| -<= 0.367 | 1,063 (17.9%) | 6 (23.1%) | 1,057 (25%) |  |
| -<= 0.502 | 1,063 (17.9%) | 7 (26.9%) | 1,056 (25%) |  |
| -<= 0.612 | 1,063 (17.9%) | 4 (15.4%) | 1,059 (25.1%) |  |
| -> 0.612 | 1,063 (17.9%) | 9 (34.6%) | 1,054 (24.9%) |  |
| Total bad debt expense for the entire hospital complex (categorized) [1,422] |  |  |  | p = 0.064 (Chi-square test) |
| -<= 1,455,721 | 1,126 (19%) | 11 (40.7%) | 1,115 (24.9%) |  |
| -<= 4,901,206 | 1,125 (19%) | 9 (33.3%) | 1,116 (24.9%) |  |
| -<= 13,793,254 | 1,125 (19%) | 5 (18.5%) | 1,120 (25%) |  |
| -> 13,793,254 | 1,126 (19%) | 2 (7.41%) | 1,124 (25.1%) |  |
| Medicare bad debt expense for the entire hospital complex (categorized) [1,689] |  |  |  | p = 0.913 (Chi-square test) |
| -<= 91,771 | 1,059 (17.9%) | 6 (28.6%) | 1,053 (25%) |  |
| -<= 272,510 | 1,059 (17.9%) | 6 (28.6%) | 1,053 (25%) |  |
| -<= 683,359 | 1,058 (17.9%) | 5 (23.8%) | 1,053 (25%) |  |
| -> 683,359 | 1,059 (17.9%) | 4 (19%) | 1,055 (25%) |  |
| Medicare allowable bad debt expense for the entire hospital complex (categorized) [1,689] |  |  |  | p = 0.913 (Chi-square test) |
| -<= 141,187 | 1,059 (17.9%) | 6 (28.6%) | 1,053 (25%) |  |
| -<= 419,246 | 1,059 (17.9%) | 6 (28.6%) | 1,053 (25%) |  |
| -<= 1,051,321 | 1,058 (17.9%) | 5 (23.8%) | 1,053 (25%) |  |
| -> 1,051,321 | 1,059 (17.9%) | 4 (19%) | 1,055 (25%) |  |
| Non Medicare and non reimbursable bad debt expense (categorized) [1,475] |  |  |  | p = 0.073 (Chi-square test) |
| -<= 1,287,139 | 1,113 (18.8%) | 11 (42.3%) | 1,102 (24.9%) |  |
| -<= 4,481,964 | 1,112 (18.8%) | 8 (30.8%) | 1,104 (25%) |  |
| -<= 12,996,729 | 1,112 (18.8%) | 5 (19.2%) | 1,107 (25%) |  |
| -> 12,996,729 | 1,112 (18.8%) | 2 (7.69%) | 1,110 (25.1%) |  |
| Total margin (categorized) [1,441] |  |  |  | p < 0.001 (Chi-square test) |
| -<= -0.021 | 1,121 (18.9%) | 21 (75%) | 1,100 (24.7%) |  |
| -<= 0.033 | 1,121 (18.9%) | 3 (10.7%) | 1,118 (25.1%) |  |
| -<= 0.094 | 1,120 (18.9%) | 3 (10.7%) | 1,117 (25.1%) |  |
| -> 0.094 | 1,121 (18.9%) | 1 (3.57%) | 1,120 (25.1%) |  |
| Medicare inpatient margin (categorized) [1,363] |  |  |  | p = 0.003 (Chi-square test) |
| -<= -0.097 | 1,141 (19.3%) | 15 (53.6%) | 1,126 (24.8%) |  |
| -<= 0.01 | 1,140 (19.2%) | 7 (25%) | 1,133 (25%) |  |
| -<= 0.048 | 1,140 (19.2%) | 2 (7.14%) | 1,138 (25.1%) |  |
| -> 0.048 | 1,140 (19.2%) | 4 (14.3%) | 1,136 (25.1%) |  |
| Medicare inpatient plus outpatient margin (categorized) [1,339] |  |  |  | p < 0.001 (Chi-square test) |
| -<= -0.113 | 1,147 (19.4%) | 17 (60.7%) | 1,130 (24.8%) |  |
| -<= 0.01 | 1,146 (19.3%) | 8 (28.6%) | 1,138 (25%) |  |
| -<= 0.018 | 1,146 (19.3%) | 1 (3.57%) | 1,145 (25.1%) |  |
| -> 0.018 | 1,146 (19.3%) | 2 (7.14%) | 1,144 (25.1%) |  |
| Operating expenses (categorized) [1,337] |  |  |  | p = 0.058 (Chi-square test) |
| -<= 28,266,941 | 1,147 (19.4%) | 12 (42.9%) | 1,135 (24.9%) |  |
| -<= 84,560,028 | 1,147 (19.4%) | 8 (28.6%) | 1,139 (25%) |  |
| -<= 242,261,893 | 1,146 (19.3%) | 6 (21.4%) | 1,140 (25%) |  |
| -> 242,261,893 | 1,147 (19.4%) | 2 (7.14%) | 1,145 (25.1%) |  |
| Operating revenues (categorized) [1,330] |  |  |  | p = 0.023 (Chi-square test) |
| -<= 26,549,815 | 1,149 (19.4%) | 12 (42.9%) | 1,137 (24.9%) |  |
| -<= 83,566,840 | 1,148 (19.4%) | 9 (32.1%) | 1,139 (24.9%) |  |
| -<= 247,066,771 | 1,148 (19.4%) | 6 (21.4%) | 1,142 (25%) |  |
| -> 247,066,771 | 1,149 (19.4%) | 1 (3.57%) | 1,148 (25.1%) |  |
| Total net expenses for cost allocation (categorized) [1,337] |  |  |  | p = 0.058 (Chi-square test) |
| -<= 25,909,408 | 1,147 (19.4%) | 12 (42.9%) | 1,135 (24.9%) |  |
| -<= 75,323,648 | 1,147 (19.4%) | 8 (28.6%) | 1,139 (25%) |  |
| -<= 219,054,008 | 1,146 (19.3%) | 6 (21.4%) | 1,140 (25%) |  |
| -> 219,054,008 | 1,147 (19.4%) | 2 (7.14%) | 1,145 (25.1%) |  |

### eTable 2

Fit statistics for confirmatory factor analysis

|  | **Statistic** |
| --- | --- |
| Number of estimated parameters in the model | 33 |
| Fit function value | 0.783 |
| Chi-squared value based on the fit function | 9,278 |
| Degrees of freedom for model | 44 |
| P-value for chi-squared value and degrees of freedom | 0 |
| Chi-squared value for baseline model | 94,093 |
| Degrees of freedom for baseline model | 55 |
| p-value for the baseline model | 0 |
| Comparative Fit Index (CFI) | 0.902 |
| Tucker-Lewis Index (TLI) | 0.877 |
| Non-Normed Fit Index (NNFI) | 0.877 |
| Relative Fit Index (RFI) | 0.877 |
| Normed Fit Index (NFI) | 0.901 |
| Parsimony Normed Fit Index (PNFI) | 0.721 |
| Incremental Fit Index (IFI) | 0.902 |
| Relative Noncentrality Index (RNI) | 0.902 |
| Logarithm of the likelihood statistic | -52,820 |
| Logarithm of the likelihood statistic for baseline model | -48,181 |
| Akaike Information Criterion (AIC) | 105,706 |
| Bayesian Information Criterion (BIC) | 105,927 |
| Total sample size | 5,924 |
| Bayesian Information Criterion Adjusted for Sample Size (BICc) | 105,822 |
| Root Mean Square Error of Approximation (RMSEA) | 0.188 (0.185, 0.191) |
| P-value associated with hypothesis of RMSEA < 0.05 | 0 |
| Root Mean Square Residual (includes means) (RMR) | 0.044 |
| Root Mean Square Residual (no means) | 0.048 |
| Standardized Root Mean Square Residual (includes means) (SRMR) | 0.035 |
| Bentler Standardized Root Mean Square Residual (includes means) | 0.035 |
| Bentler Standardized Root Mean Square Residual (no means) | 0.038 |
| Bollen Standardized Root Mean Square Residual (includes means) | 0.037 |
| Bollen Standardized Root Mean Square Residual (no means) | 0.04 |
| MPlus Standardized Root Mean Square Residual (includes means) | 0.035 |
| MPlus Standardized Root Mean Square Residual (no means) | 0.037 |
| Critical n for alpha = 0.05 | 39.6 |
| Critical n for alpha = 0.01 | 44.9 |
| Goodness-of-Fit Index (GFI) | 0.828 |
| Adjusted Goodness-of-Fit Index | 0.7 |
| Parsimony Goodness-of-Fit Index (PGFI) | 0.473 |
| McDonald Fit Index (MFI) | 0.459 |
| Expected Cross-Validation Index | 1.58 |
