## Supplementary material for "Psychometric development and validation of the Hospital Resilience Index": eFigures legends

### Legends to Supplemental Figures

### eFigure 1

Correlation graph between selected variables. The coefficients are considered different from zero (significant), with 95% confidence assuming a normal distribution, when the calculated intervals did not include zero, that is, p-value>0.05. Black X’s above the correlation coefficient value mean that the correlation is not significant.

### eFigure 2

Exploratory factor analysis diagram.

### eFigure 3a

Contribution of each item from Hospital Resilience Index to the predicted probability of closure for three hospitals that did close in 2019.

### eFigure 3b

Contribution of each item from the Hospital Resilience Index to the predicted probability of closure for three hospitals that did not close in 2019.

### eFigure 4a

Contribution of each item from the Hospital Resilience Index to the predicted probability of closure for three hospitals that closed in 2020.

**eFigure 4b**

Contribution of each item from the Hospital Resilience Index to the predicted probability of closure for three hospitals that did not close in 2020.

**eFigure 5**

### Evaluation of hospital resilience index (HRI) for Venice Regional Bayfront Health over three consecutive years

### eFigure 6

Evaluation of HRI by year for hospitals that closed in 2020 versus hospitals that did not close in that year.
