## Supplementary figures and images for "Psychometric development and validation of the Hospital Resilience Index"

### eFigure 1

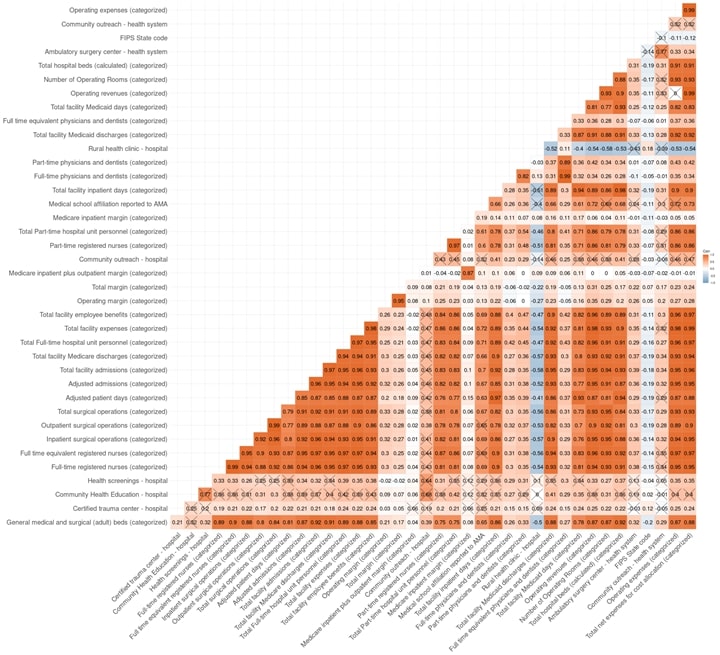

### eFigure 2

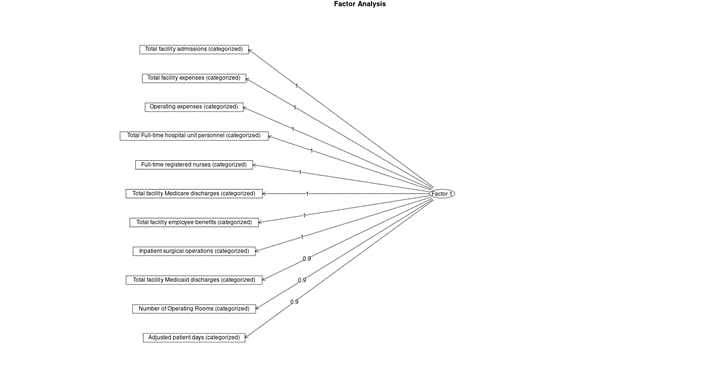

### eFigure 3a

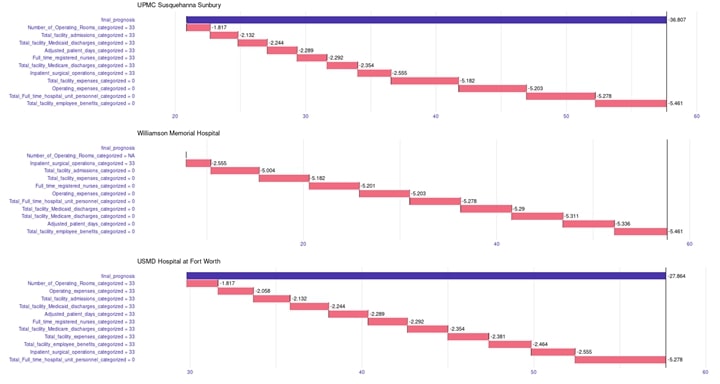

### eFigure 3b

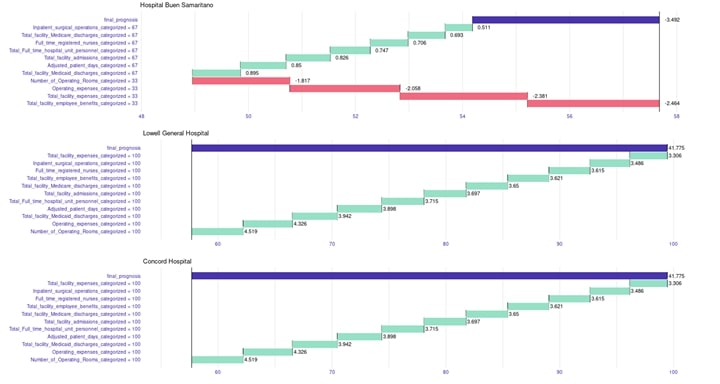

### eFigure 4a

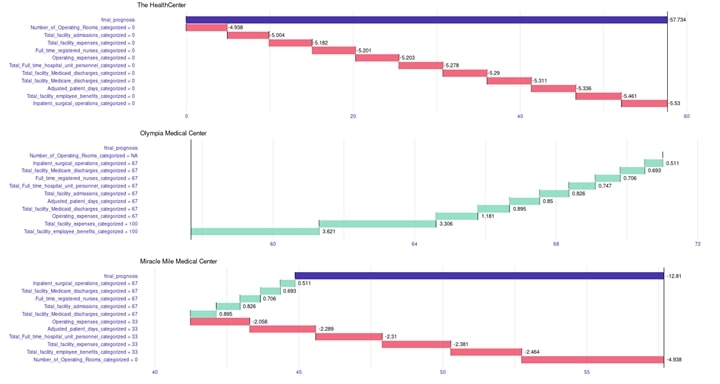

### eFigure 4b

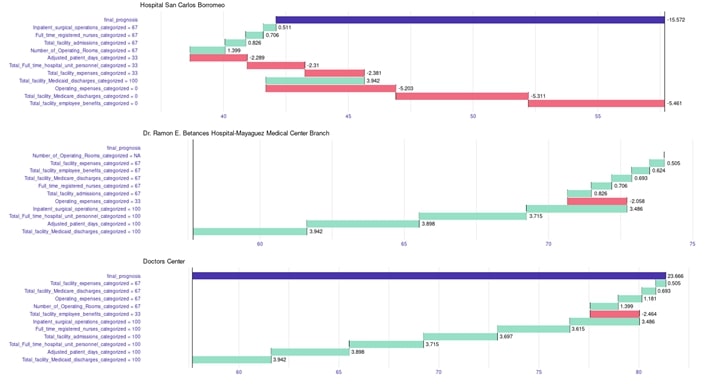

### eFigure 5

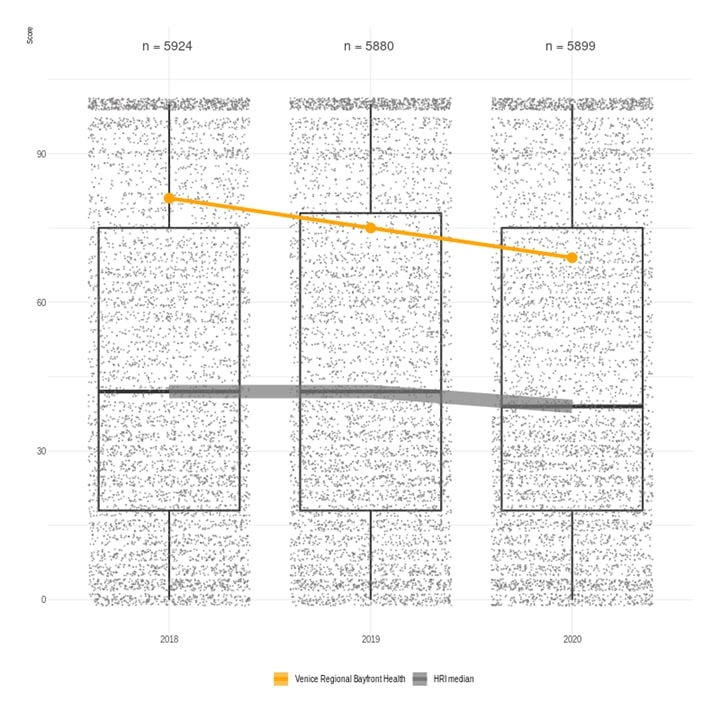

### eFigure 6

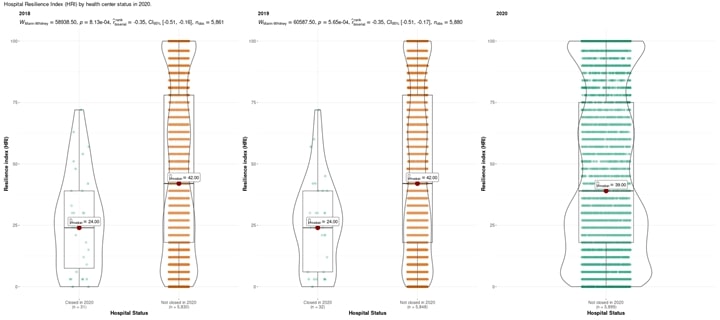
